# Loss of coactosin-like F-actin binding protein 1 (Cotl1) decreases platelet-mediated osteoclastogenesis and causes osteopetrosis phenotypes in mouse

**DOI:** 10.1101/2023.12.20.23300337

**Authors:** Eunkuk Park, Seung-Hee Yun, Hyun-Seok Jin, Chang-Gun Lee, So-Hyun Lee, Seok-Yong Choi, Hyun Goo Woo, Ji Eun Lim, Bermseok Oh, Seon-Yong Jeong

## Abstract

**BACKGROUNDS:** Osteopetrosis, a rare skeletal disease, is characterized by an increased bone mass resulting from impaired bone remodeling process. Platelet is the major bone-healing blood component involved in the regulation of bone resorption, particularly in the removal of compromised bones. Several actin-associated proteins contribute to the orchestration of actin ring formation in osteoclasts closely related to bone resorption. However, the role of coactosin-like F-actin binding protein 1 (Cotl1) in actin ring formation and platelet-mediated bone resorption mechanisms remains unclear.

**METHODS:** Whole-mount *in situ* RNA hybridization was performed to detect cotl1 expression pattern in zebrafish. *cotl1* gene knockdown zebrafish using morpholino oligonucleotides and platelet marker-expressing transgenic zebrafish were investigated for finding the phenotypic clues. *Cotl1* knockout (*Cotl1*^-/-^) mice were generated using *Cre/loxP* recombination systems. *In silico* network analysis of the differentially expressed genes between bone marrow samples of wild type and *Cotl1*^-/-^ mice was conducted. Primary-cultured monocytes from *Cotl1*^-/-^ mice were examined for osteoclast differentiation and mRNA and protein expression patterns. *Cotl1*^-/-^ mice underwent hematological examination and bone phenotype assessments including micro-CT, bone density, histology, immunohistochemistry, electron microscopy, and mechanical testing. Genetic association of SNPs in human *COTL1* gene with estimated bone mineral density was analyzed.

**RESULTS:** Zebrafish *cotl1* mRNA was highly expressed in the caudal hematopoietic tissue region. Knockdown of *cotl1* in zebrafish embryos decreased the expression of *c-myb*, a marker of hematopoietic stem cells (HSCs). Notably, the platelet receptor CD41 was reduced in the HSCs of *cotl1-*depleted zebrafish and *Cotl1*^-/-^ mice showed reduced platelet production with platelet surface markers of CD41 and CD61. Significantly reduced osteoclast differentiation and bone resorption pit, and impaired actin ring formation were observed in the primary myocytes from *Cotl1*^-/-^ mice. Structural and histological analyses of the femur revealed sclerotic bone phenotypes in *Cotl1*^-/-^ mice. Mechanical assessment of *Cotl1*^-/-^ mouse femoral bones revealed osteopetrotic phenotypes. Association analysis of genetic variants in *COTL1* gene in subjects from the UK Biobank suggested that *COTL1* is susceptible to bone density in humans.

**CONCLUSIONS:** Our results provide insights into the role of Cotl1 in platelet-mediated osteoclastogenesis and the novel finding that the loss of *Cotl1*^-/-^ mice causes osteopetrosis phenotypes.

**Clinical Perspective:** **What Is New?**

- Deficiency of Cotl1 decreased platelet production in zebrafish and mice.
- Absence of Cotl1 disrupted the actin ring formation which is crucial for osteoclast differentiation in bone remodeling process.
- *Cotl1* knockout mice displayed sclerotic bone phenotypes and increased bone density that are representative characteristics of osteopetrosis.
- Genetic variants in *COTL1* gene in subjects from the UK Biobank are significantly associated with bone density.

**What Are the Clinical Implications?**

- The current findings suggest that Cotl1 plays a fundamental role in platelet production-mediated osteoclastogenesis during bone remodeling, providing valuable insights into novel strategies for bone health maintenance.
- Cotl1 may be a promising target for novel therapeutic strategies for the treatment and/or prevention of impaired osteoclastogenesis-mediated bone diseases such as osteopetrosis and osteoporosis.

## INTRODUCTION

Osteopetrosis is a genetic skeletal disease primarily caused by abnormal osteoclastogenesis^1^. Autosomal dominant osteopetrosis is the most common disorder, with an incidence of 1 in 20,000 births, whereas autosomal recessive osteopetrosis has an incidence of 1 in 250,000 births and is categorized as a rare disease^2^. Osteopetrosis is characterized by an excessive increase in bone density and susceptibility to fractures, resulting in pain, swelling, and stiffness^1^. Owing to the increased bone mass, osteopetrosis can cause abnormal syndromes such as macrocephaly, craniofacial dysostosis, and bone marrow failure syndrome^2^. Interrupted alteration of the bone structure is prompted by imbalanced bone remodeling, a metabolic process that maintains healthy bone^3^.

Bone remodeling is a physiological process in which old or damaged bone tissue is resorbed by osteoclasts and new bone is synthesized by osteoblasts^3^. The hematopoietic lineage is characterized by various interactions between mesenchymal osteoblasts and monocytic lineage^4^. Thus, abnormal hematopoiesis from an insufficient bone marrow cavity can cause unbalanced bone remodeling. Osteoblast differentiation from mesenchymal stem progenitor cells is responsible for bone synthesis^4^ and is regulated by secreting osteoclastogenesis-related cytokines, including receptor activator of nuclear factor kappa-light-chain-enhancer of activated B cells (NF-κB) ligand (RANKL), macrophage colony-stimulating factor (M-CSF), and osteoclast differentiation inhibiting factors such as osteoprotegerin (OPG)^5^. Osteoclasts are giant multinucleated cells that originate from the monocyte/macrophage lineage in the bone marrow and play a critical role in the regulation of bone resorption^5^. Bone marrow-derived monocytes/macrophages require two critical cytokines, RANKL and M-CSF, for differentiation into mature osteoclasts^5^. M-CSF promotes the proliferation and survival of osteoclast progenitors that express the receptor activator of NF-κB (RANK)^6^. Binding of RANK to RANKL stimulates the master transcription factor of osteoclasts, resulting in the formation of mature multinuclear osteoclasts^6, 7^.

During bone resorption, mature osteoclasts attach to the bone extracellular matrix and polarize to form a ruffled border and an actin ring^8^. The ruffled border serves as a gateway for protons and lysosomal proteases^9^. Protons cause acidification to provide an optimal pH (4.0) for lysosomal protease activity, inducing the digestion of damaged or old bone minerals^10^. Lysosomal proteases undergo proteolysis to degrade the organic matrix of the bone, which is mainly composed of type I collagen fibers^11^. The actin ring seals the ruffle border to maintain acidification of the resorption pit and high levels of lysosomal protease^9^. The actin ring is organized into podosomes and actin cytoskeletal structures, which mediate the adhesion of osteoclasts to the bone plate^12^. In several cells including macrophages and dendritic cells, podosomes exist in individual dot patterns^13^, whereas podosomes in a ring pattern (actin rings) are found only in mature osteoclasts^14^. Cytoskeleton-related proteins involved in the formation of actin rings have been highlighted as potential regulators of bone remodeling^15, 16^. Actin polymerization and depolymerization are regulated by actin-depolymerizing factor homology (ADF-H) domain-containing family proteins such as cofilin^17^. Cofilin, which regulates actin network homeostasis^17^, interacts with cortactin and regulates the osteoclast actin ring^15^. Therefore, actin-associated proteins play an important role in the regulation of actin ring formation related to bone resorption.

Coactosin-like protein 1 (Cotl1), an ADF-H domain-containing protein, is tightly associated with F-actin bundles and promotes the assembly of protrusive actin filament arrays at the leading edges of growth cones for axon guidance^18^. Furthermore, Cotl1 competes with cofilin to bind F-actin in B cells^19^. However, unlike cofilin, the role of Cotl1 in actin ring formation in osteoclasts and bone resorption has not yet been studied.

In this study, we aimed to explore how Cotl1 is involved in actin ring formation in osteoclasts which is closely related to osteoclastogenesis and further bone resorption during bone remodeling. First, we performed knockdown experiments of *cotl1* in zebrafish to identify phenotypic clues during embryonic development. Next, we established *Cotl1* knockout (*Cotl1*^-/-^) mice and investigated the effects of *Cotl1* loss on platelet production and osteoclast differentiation in primary cultured monocytes. Based on the *in vitro* and *in silico* pathway analysis results, we observed bone features in male and female *Cotl1*^-/-^ mice. This study provides insights into the role of Cotl1 in platelet-medicated osteoclastogenesis and the novel finding that the loss of *Cotl1*^-/-^ mice causes osteopetrosis phenotypes.

## METHODS

### Zebrafish studies

Wild-type (WT) zebrafish (AB strain) were obtained from the Zebrafish International Resource Centre (Eugene, OR, USA) and staged in hours post-fertilization (hpf) or days post-fertilization (dpf). *In situ* hybridization was performed using *cotl1* or *c-myb* riboprobes. *cotl1* morpholino oligonucleotides (6 ng) or *cotl1* mRNA (70 pg) were injected into one-cell stage embryos. The injected embryos were incubated until 2 dpf and probed with *c-myb* riboprobes. Images of platelet *CD41 – GFP* transgenic zebrafish were captured using a confocal microscope.

### Knockout mice studies

*Cotl1* heterozygote mice (*Cotl1*^+/-^) in the C57BL/6 background were obtained from the European Mouse Mutant Archive. *Cotl1-*knockout (*Cotl*^-/-^) mice were generated using *Cre/loxP* recombination systems targeting the third exon of the *Cotl1* gene. To mimic osteoporosis in a murine model, surgical dissection of the ovaries was performed in 8-week-old female mice. All animal research procedures were approved by the Animal Care and Use Committee of the Ajou University School of Medicine (IACUC No. 2021-0016), and the experiments were conducted in accordance with the institutional guidelines established by the committee. For primary culture of osteoblasts from both WT and *Cotl1^-/-^*mice, calvaria were dissected from 5-day-old mouse pups, and osteoblastic cells were isolated. For primary culture of osteoclasts, bone marrow cells were flushed from the femoral bones of 6-week-old mice.

### Association studies in the UK Biobank dataset

The UK Biobank dataset, a population-based cohort that recruited more than 488,133 individuals aged 40-69 years between 2006-2010, was used for association study. UK Biobank was granted ethical approval to collect participant data by the North West Multicenter Research Ethics Committee, the National Information Governance Board for Health & Social Care, and the Community Health Index Advisory Group (21/NW/0157). This study received approval to access the UK Biobank genetic and phenotypic data under application number 83990. After filtration process, total 357,916 unrelated white British subjects were selected. Association analysis of total 6,047 single nucleotide polymorphisms (SNPs) in *COTL1* gene with estimated bone mineral density (eBMD) was performed using PLINK version 1.90. All association tests were based on the additive genetic model, and probability value (*p*-value) were adjusted for multiple tests using the Bonferroni-corrected significance level.

### Statistical analysis

Data in the bar graphs are expressed as the mean ± standard error of the mean calculated using GraphPad Prism 9.0 software (San Diego, CA, USA). Comparisons between multiple groups were performed by one-way analysis of variance with Tukey’s honest significant difference post-hoc test, while comparisons between two groups were performed by Student’s t-test using the professional Statistical Package for the Social Sciences (SPSS 11.0 for Windows, SPSS Inc. Chicago, IL, USA); p < 0.05 was considered statistically significant.

## RESULTS

### Effect of *cotl1* knockdown in zebrafish

We first examined the expression pattern of *cotl1* during embryo development using whole-mount *in situ* RNA hybridization. Embryos were treated with 1-phenyl-2-thiourea to suppress melanin synthesis. *cotl1* mRNAs were highly expressed in the aorta-gonad-mesonephros at 40 h post-fertilization, and in the caudal hematopoietic tissue region at 3- and 4-days post-fertilization (Figure S1a). Next, we investigated the effects of *cotl1* knockdown by injecting morpholino RNAs into the zebrafish embryos. Successful depletion of wild-type *cotl1* was achieved with morpholino oligonucleotides targeting the exon 2–intron 2 junction site of *cotl1* in one-cell-stage embryos (Figure S1b, c). As the predominant mRNA expression of *cotl1* was found in hematopoietic tissue, we observed hematopoietic stem cells (HSCs) in zebrafish. *In situ* hybridization of *cotl1* morphants revealed significantly decreased expression of *c-myb*, an HSC marker (Fig. 1a, b; Figure S1d). Injection of *cotl1* mRNA into *cotl1* morphants recovered the hematopoietic stem cell numbers (Fig. 1a, b). Furthermore, transgenic platelet glycoprotein IIb integrin α 2b (*CD41*)*-*green fluorescent protein (*GFP*)-expressing cell number and movement were decreased in *cotl1* morphant zebrafish compared to wild-type (WT) control zebrafish (Fig. 1c; Figure S1e). These results indicate that knockdown of *cotl1* may influence hematopoiesis, including platelet production, in zebrafish.

**Figure 1.**
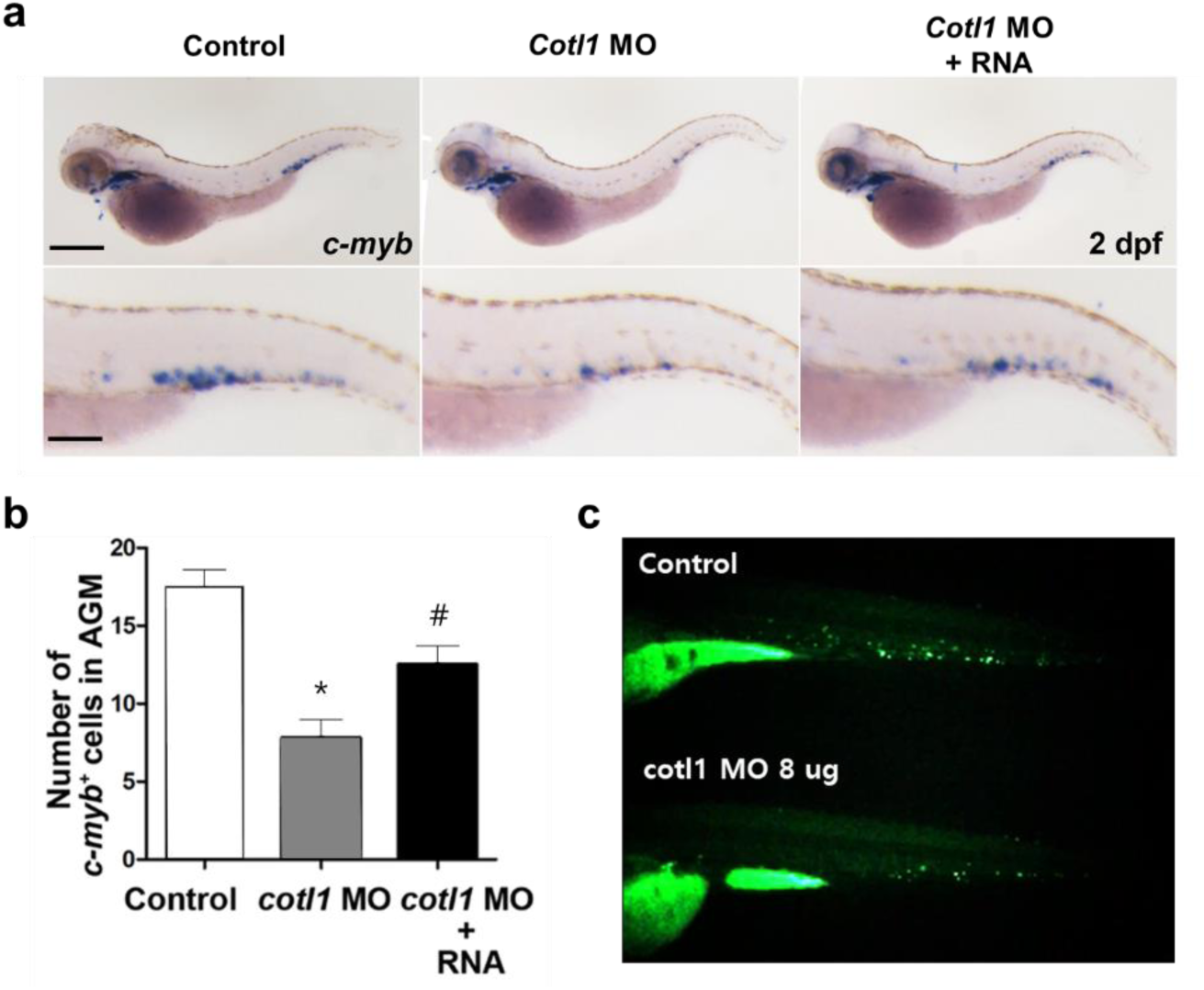
Effects of *cotl1* on the hematopoietic systems in zebrafish. **a,** One-cell-stage zebrafish embryos were injected with either control morpholino oligonucleotides (MOs) or *cotl1* MOs, fixed at 2 days post-fertilization (dpf), probed with *c-myb* riboprobes (a marker of hematopoietic stem cells), and imaged under a stereomicroscope. For the rescue experiment, one-cell-stage zebrafish embryos were sequentially injected with *cotl1* MO and zebrafish *cotl1* RNA (*cotl1* MO + RNA), fixed at 2 dpf, and subjected to *in situ* hybridization with *c-myb* riboprobes. Lower panels show magnifications of the aorta-gonad-mesonephros (AGM) region of the respective upper panel images. Scale bars: upper panel = 250 µm; lower panel = 100 µm. **b,** Number of *c-myb*+ cells in the AGM in each group. \**p* < 0.05 vs. Control and ^#^*p* < 0.05 vs. *cotl1* MO (one-way analysis of variance with Tukey’s multiple comparison test). **c,** Photos of transgenic *CD41–GFP* zebrafish captured at 3 dpf under a stereomicroscope.

### Effects of *Cotl1* loss on platelet levels and bone features in mice

CD41, a platelet receptor, is a reliable biomarker for the early developmental stages of hematopoiesis and multipotent primitive HSCs play an important role in the production of blood platelets^20^. Based on the results of transgenic *CD41–GFP*-expressing *cotl1* knockdown zebrafish, we further explored the inhibitory effects of *Cotl1* on platelet production in *Cotl1* knockout mice. *Cotl1*^-/-^ mice were generated by *Cre/loxP* recombination system targeting the third exon of *Cotl1* (Figure S2a). No abnormality in morphology, behavior, dietary intake, lifespan, reproduction was observed in *Cotl1*^-/-^ mice until the 10th filial generation (data not shown). Hematological examination showed no differences in blood parameters between WT and *Cotl1*^-/-^ mice at 7, 12, and 18-week of ages; however, at 24-week of age, platelet production in both male and female *Cotl1*^-/-^ mice was significantly reduced compared to that in WT mice (Fig. 2a; Figure S2b). Consistently, the levels of the platelet surface markers CD41 and CD61 (platelet GPIIIa, integrin β3) were decreased in 24-week-old *Cotl1*^-/-^ mice (Fig. 2a; Figure S2c). These findings indicated a critical role for *Cotl1* in blood platelet production.

**Figure 2.**
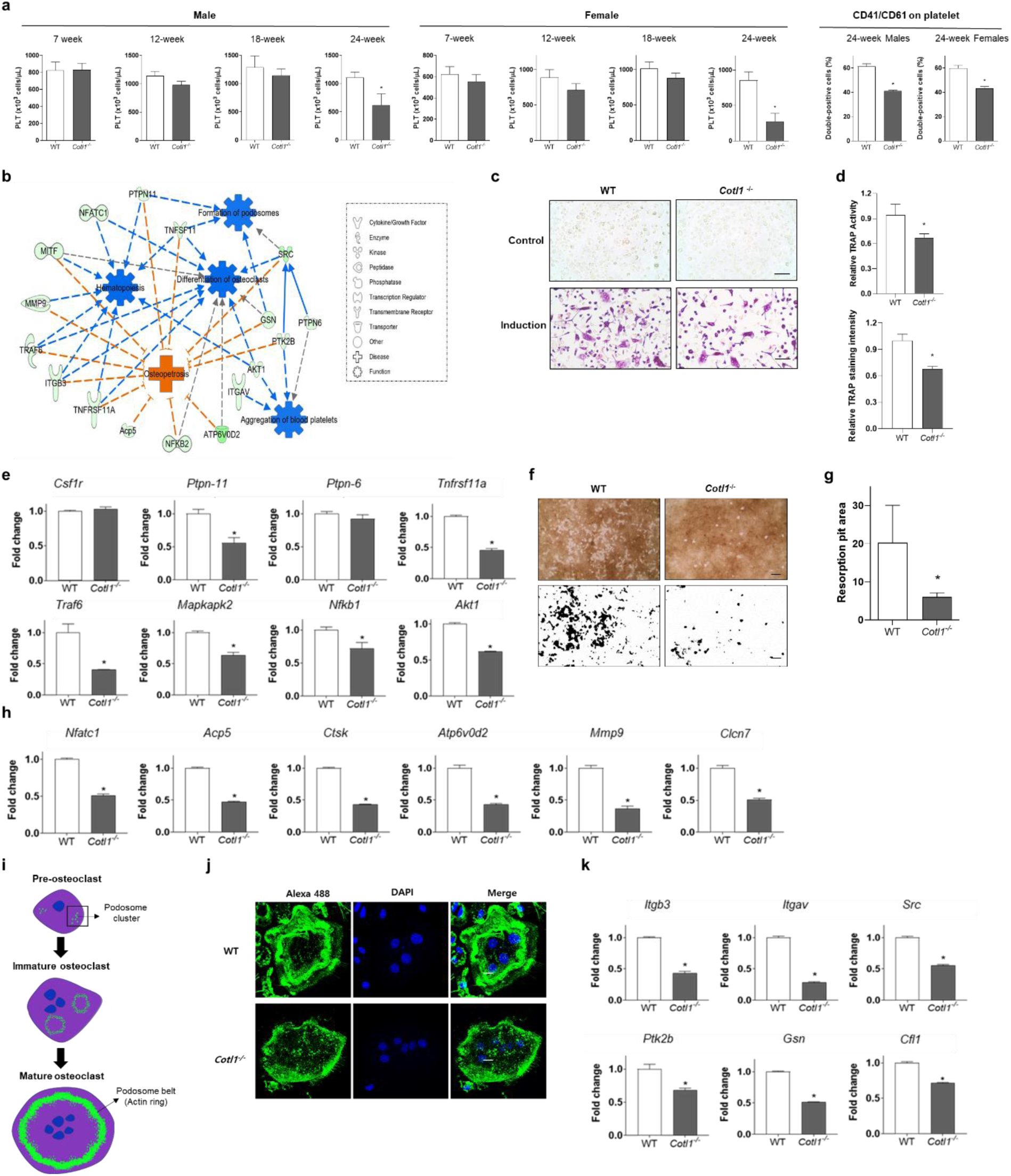
Effects of *Cotl1* knockout on platelet production and osteoclastogenesis. **a,** Hematological analysis of platelets counted at 7, 12, 18, and 24 weeks in male and female wild-type (WT) and *Cotl1^-/-^* mice. Expressions of specific platelet membrane glycoproteins CD41 (platelet glycoprotein IIb, integrin α2b) and CD61 (platelet GPIIIa, integrin β3) on platelet (PTL) were analyzed by flow cytometry. **b,** *In silico* network analysis of the differentially expressed genes (DEGs) in between bone marrow cells of WT and *Cotl1*^-/-^ mice. DEGs between WT and *Cotl1*^-/-^ mice are associated with gene function keywords, including ‘Osteopetrosis’, ‘Hematopoiesis’, ‘Formation of podosomes’, and ‘Differentiation of osteoclasts’. The pathway networks of the connected molecules were formatted using Ingenuity Pathway Analysis software with the QIAGEN Knowledge Base. Solid lines indicate direct interaction and dashed lines indicate indirect interaction. **c,** Osteoclast differentiation evaluated by tartrate-resistant acid phosphatase (TRAP) staining. The primary monocytes of WT and *Cotl1*^-/-^ mice were differentiated by administration of M-CSF and 100 ng/mL of RANKL. Scale bar, 100 μm. **d,** Relative TRAP activity and TRAP-positive cell intensity analyzed by ImageJ software. **e,** mRNA expression levels of osteoclastogenesis markers determined using quantitative reverse-transcription (RT)-PCR: colony-stimulating factor 1 receptor (*Csf1r*), protein tyrosine phosphatase non-receptor type 6 and 11 (*Ptpn-6 and 11*), TNF receptor superfamily member 11a (RANK, encoded by *Tnfrsf11a*), TNF receptor-associated factor 6 (*Traf6*), MAP kinase-activated protein kinase 2 (*Mapkapk2*), nuclear factor of kappa light polypeptide (*Nfkb*), and AKT serine/threonine kinase 1 (*Akt1*). For the examination of bone resorption, cells were removed by adding 5% sodium hypochlorite after 5 days of incubation in the osteoclast induction medium. **f, g,** Resorption pit images captured under a light microscope and resorption pit area calculated using ImageJ software. **h**, mRNA expression levels of the bone resorption markers determined using quantitative RT-PCR: nuclear factor of activated T cells, cytoplasmic, calcineurin dependent 1 (*Nfatc1*), acid phosphatase 5, tartrate resistant (TRAP, encoded by *Acp5*), cathepsin K (*Ctsk*), ATPase H+ transporting V0 subunit D2 (*Atpv0d2*), matrix metalloproteinase-9 (*Mmp9*), and chloride voltage-gated channel 7 (*Clcn7*). **i,** Actin ring structure formation observed during osteoclast differentiation **j.** Representative images of differentiated osteoclasts isolated from WT and *Cotl1*^-/-^ mice; F-actin is stained with Alexa-488 (green) and nuclei are stained with DAPI (blue). Scale bars, 20 μm. **k,** mRNA expression levels of the cytoskeleton markers determined using quantitative RT-PCR: integrin β3 (*Itgb3*), integrin αV (*Itgav*), SRC proto-oncogene (*Src*), PTK2 protein tyrosine kinase 2 beta (*Ptk2b*), gelsolin (*Gsn*), and cofilin 1 (*Cfl1*) in WT and *Cotl1*^-/-^ mouse primary osteoclasts. Relative gene expression levels were normalized by mouse *Hprt1* expression. \**p* < 0.05 vs. WT (Student’s *t*-test).

Platelets are important regenerative factors that regulate hemostasis, inflammation, and bone remodeling^21^. To identify differentially expressed genes (DEGs), RNA sequencing of bone marrow isolated from WT and *Cotl1*^-/-^ mice was performed using a next-generation sequencer. Many differentially expressed genes (DEGs) were identified (data not shown). *In silico* pathway network analysis of the DEG data was performed using the Path Explorer tool in Ingenuity Pathway Analysis software. Gene networks of DEGs showed close associations with gene functional terminologies of ‘podosome formation,’ ‘hematopoiesis,’ ‘platelet aggregation,’ ‘osteoclast differentiation’ and ‘osteopetrosis’ (Fig. 2b), suggesting that *Cotl1* may be involved in bone homeostasis as well as actin ring formation and hematopoiesis. Therefore, we focused on identifying abnormalities in the bone features of *Cotl1*^-/-^ mice. Alkaline phosphatase (ALP) activity test and ALP staining of primary cultured pre-osteoblasts isolated from the mouse calvaria showed no difference in osteoblast differentiation between WT and *Cotl1*^-/-^ mice (Figure S3a–c). However, tartrate-resistant acid phosphatase (TRAP) activity test of primary bone marrow-derived monocytes isolated from the femurs showed a significantly decreased number of differentiated multinuclear osteoclasts in *Cotl1*^-/-^ mice compared to WT mice (Fig. 2c, d; Figure S4a–c). Furthermore, Cotl1 protein expression was increased by the induction of osteoclast differentiation (Figure S4d). Similarly, decreased mRNA expression levels of osteoclastogenesis markers were found in *Cotl1*^-/-^ mice (Fig. 2e; Figure S4e).

A bone resorption assay of osteoclasts revealed that the resorption pit area was significantly decreased in *Cotl1*^-/-^ mice compared to WT mice (Fig. 2f, g; Figure S5a, b). Furthermore, the mRNA expression levels of bone resorption markers were significantly decreased in *Cotl1*^-/-^ mice than WT mice (Fig. 2h; Figure S5c). Actin rings forming podosome assemblies were observed only in mature osteoclasts (Fig. 2i). Therefore, we investigated whether loss of *Cotl1* affects cytoskeletal formation during osteoclast differentiation. The formation of actin filaments was not detected in immature osteoclasts 3 days after induction in either WT or *Cotl1*^-/-^ mice (Figure S6a). Actin ring formation in mature osteoclasts of WT mice was completed after 5 days, but was not observed in mature osteoclasts of *Cotl1*^-/-^ mice (Fig. 2j; Figure S6b). Impaired actin ring formation due to *Cotl1* loss was further demonstrated by a significant reduction in the mRNA expression of cytoskeletal regulatory markers (Fig. 2k; Figure S6c). These results suggested an important role of Cotl1 in platelet production in the bone marrow and actin ring formation-mediated osteoclastogenesis during bone remodeling.

### Structural and histological analyses of bone in the *Cotl1^-/-^* male mice

Next, we examined the effects of *Cotl1* knockout on bone features in male mice. Three-dimensional, sagittal, and axial micro-computed tomography (micro-CT) images of the femoral bones in *Cotl1*^-/-^ male mice showed increased structural bone density compared to that in WT male mice (Fig. 3a; Figure S7a–c). Furthermore, bone mineral density (BMD) was significantly higher in *Cotl1*^-/-^ mice than WT mice from four-week old to 24-week old ages (Fig. 3b). Micro-CT of the microstructural bone properties showed increased structural parameters of the femoral bones, including BMD, trabecular bone thickness (Tb.Th), trabecular space (Tb.sp), trabecular number (Tb.N), and bone volume (BV/TV) in *Cotl1^-/-^*mice (Fig. 3c). Fewer TRAP-positive cells in the femur was observed in *Cotl1^-/-^* mice, but there was no difference in the number of ALP-stained cells between WT and *Cotl1*^-/-^ mice (Fig. 3d, e). Consistently, the results of scanning electron microscopy (SEM) combined with TRAP staining demonstrated reduced osteoclasts in *Cotl1^-/-^* mice compared to WT mice at 10- and 18-week of ages (Fig. 3f; Figure S8a, b). The protein levels of bone remodeling-associated factors in the blood plasma showed increased OPG and decreased RANKL, resulting in an increase in the OPG/RANKL ratio in *Cotl1*^-/-^ male mice (Fig. 3g). These results indicated that knockout of *Cotl1* in male mice caused osteopetrosis phenotypes.

**Figure 3.**
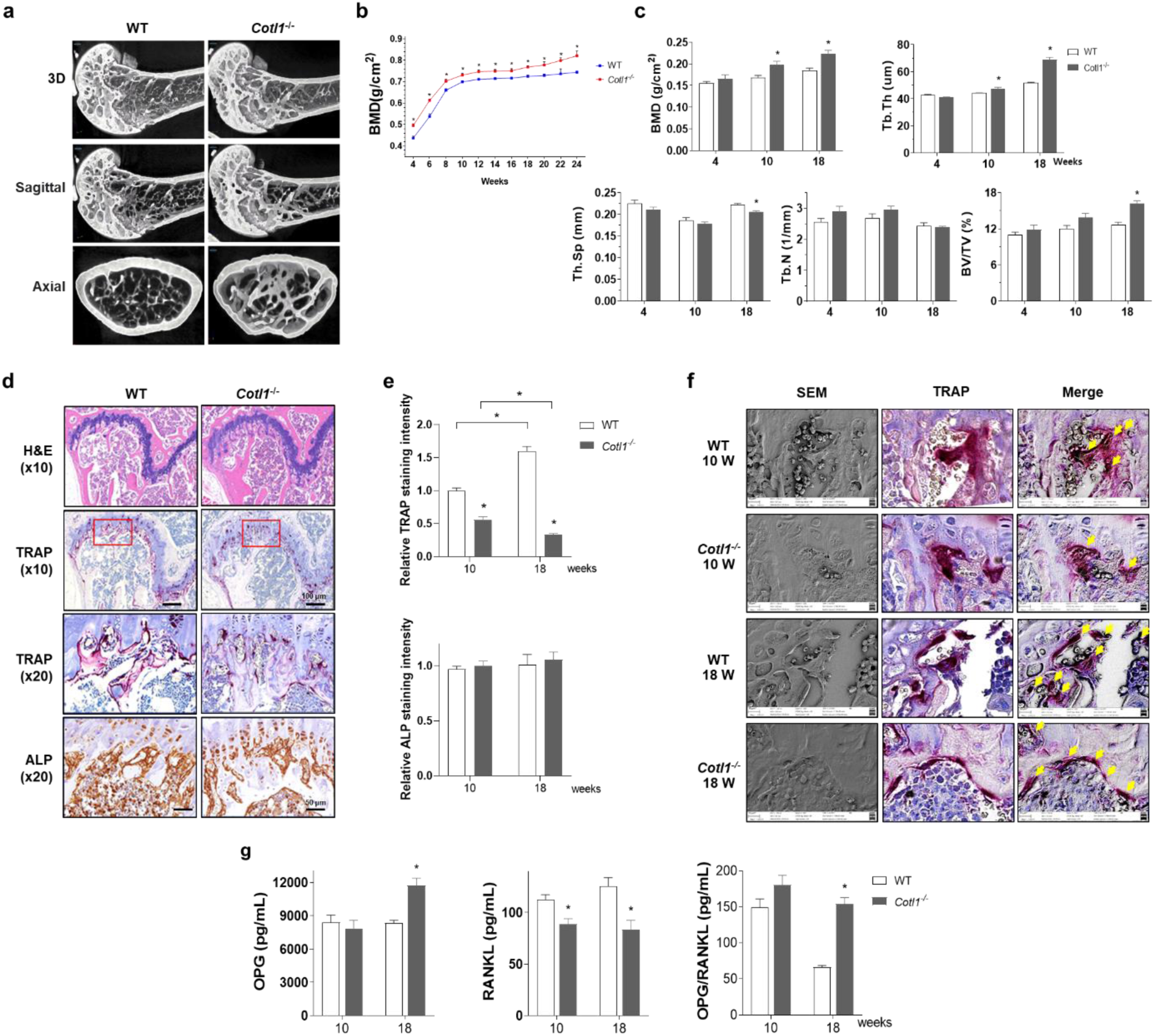
Effects of *Cotl1* knockout on the bone features in male mice. **a.** Representative images of micro-computed tomography (micro-CT) of the femurs of 18-week-old male mice. **b, c**, Bone mineral density (BMD) of the right femur in wild-type (WT) and *Cotl1*^-/-^ mice from 4–24 weeks. Quantification of bone structural parameters of micro-CT from 4-, 10-, and 18-week-old male mice, including BMD, bone volume (BV/TV), trabecular bone thickness (Tb.Th), trabecular space (Tb, sp), and trabecular number (Tb.N). **d.** Representative images of hematoxylin and eosin (H&E), tartrate-resistant acid phosphatase (TRAP), and alkaline phosphatase (ALP) immunohistochemistry staining. **e.** Relative TRAP and ALP staining intensity in the femurs of male mice. **f.** Representative images of scanning electron microscopy and TRAP immunohistochemistry staining at 10- and 18-week-old ages in WT and *Cotl1*^-/-^ male mice. **g,** Blood plasma levels of bone remodeling-associated factors, including osteoprotegerin (OPG) and receptor activator of NF-κB ligand (RANKL). \**p* < 0.05 vs. WT (Student’s t-test).

### Structural and histological analyses of bones in the sham and ovariectomized (OVX) *Cotl1*^-/-^ female mice

We further investigated the effects of *Cotl1* knockout on bone features in unmanipulated sham controls (non-surgical treatment) and OVX-induced osteoporotic female mice. Micro-CT images and BMD measurements revealed that *Cotl1^-/-^* female sham and OVX mice showed increased structural bone density and BMD compared to WT female sham and OVX mice (Fig. 4a-d; Figure S9a). OVX induced a significant decrease in microstructural properties and increased osteoclastic activity in the femurs of OVX-induced WT mice. However, improved structural bone density and augmented BMD were observed throughout the experimental period in OVX-treated female *Cotl1^-/-^* mice (Fig. 4c, d; Figure S9a). Furthermore, structural trabecular properties, including BMD, BV/TV, Tb.Th, and Th.Sp, were increased by *Cotl1* loss in both control and OVX-induced mice (Fig. 4e). Histological analyses of TRAP staining showed significantly reduced osteoclast differentiation in *Cotl1^-/-^*mice compared to WT mice, but there was no difference in ALP staining between WT and *Cotl1*^-/-^ mice (Fig. 4f, g). SEM images merged with TRAP staining showed clear differences in osteoclastogenesis between control and OVX-induced WT and *Cotl1*^-/-^ mice (Fig. 4h; Figure S9b). Similar bone phenotypes were observed in 24-week-old *Cotl1*^-/-^ female mice (Figure S10a–c). These results demonstrate that the loss of *Cotl1* in mice results in osteopetrosis phenotypes caused by inhibited osteoclastogenesis.

**Figure 4.**
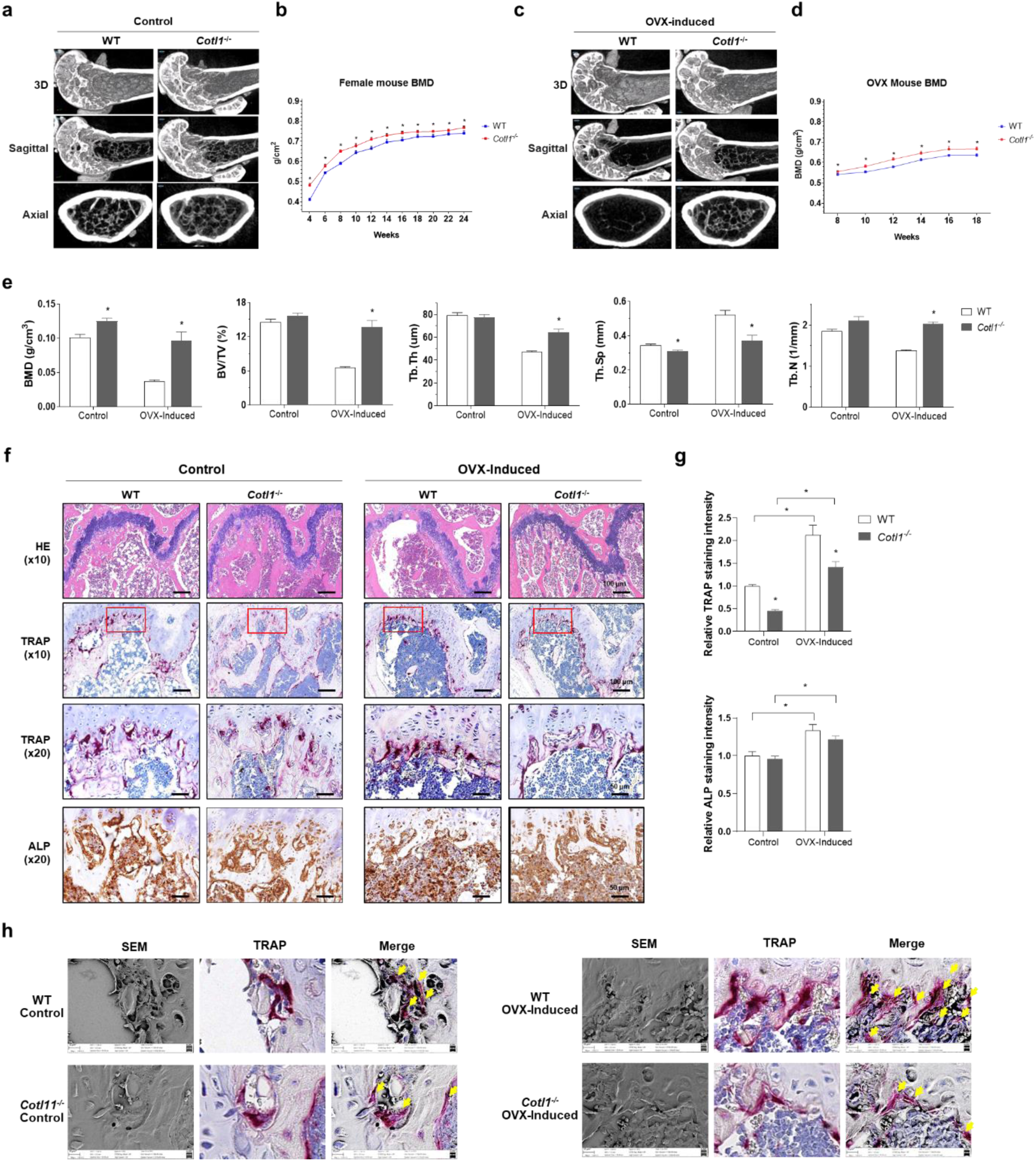
Effects of *Cotl1* knockout on bone features in the sham and ovariectomized (OVX) female mice. **a, c,** Representative images of micro-computed tomography of the femurs of 18-week-old female unmanipulated sham (Control) and ovariectomized-induced (OVX) mice. **b, d**, Bone mineral density (BMD) of the right femur in wild-type (WT) and *Cotl1*^-/-^ mice from 4–24 weeks and in OVX-induced WT and *Cotl1*^-/-^ mice from 8–24 weeks. **e,** Quantification of bone structural parameters from micro-CT in control and OVX-induced WT and *Cotl1*^-/-^ mice at 18-week-old age. **f.** Histological images of hematoxylin and eosin (H&E), tartrate-resistant acid phosphatase (TRAP), and alkaline phosphatase (ALP) staining. **g,** Relative TRAP and ALP staining intensity in the femurs of control and OVX-induced WT and *Cotl1*^-/-^ mice at 18-week-old age. **h**, Representative images of scanning electron microscopy and TRAP immunohistochemistry staining at 18-week-old mice. \**p* < 0.05 vs. WT (Student’s *t*-test).

### Mechanical assessment of the femoral bone in *Cotl1*^-/-^ mice

Finally, we compared the mechanical properties of the femoral bones between WT and *Cotl1*^-/-^ mice. The femur stiffness was measured using a three-point bending test, in which the slope of the initial lines of the load–displacement curve represented the elastic deformation when the structure was under load (Fig. 5a). The yield load was defined as the point at which the curve deviated from the linear curve, indicating irreversible damage (Fig. 5b). The post-yield displacement (PYD) was determined as the displacement from the yield point to the fracture point, indicating the mechanical characteristics of brittleness (absence of PYD or very low PYD) or ductility (opposite of brittleness) (Fig. 5b). The work to fracture was analyzed by the area of the load-displacement curve until fracture (Fig. 5b), implying an overall resistance to fracture^22^. Under similar maximum loads, the displacement from the yield point to the fracture point in *Cotl1*^-/-^ mice was shorter than that in WT mice, indicating that the femoral bones of *Cotl1*^-/-^ mice were at a high risk of fracture (Fig. 5c). Furthermore, the femoral bone of *Cotl1*^-/-^ mice appeared to have significantly increased stiffness and yield load, and significantly decreased PYD and work-to-fracture compared to those of WT mice (Fig. 5d). These results demonstrated that *Cotl1*^-/-^ mouse femoral bone presented a marble bone phenotype.

**Figure 5.**
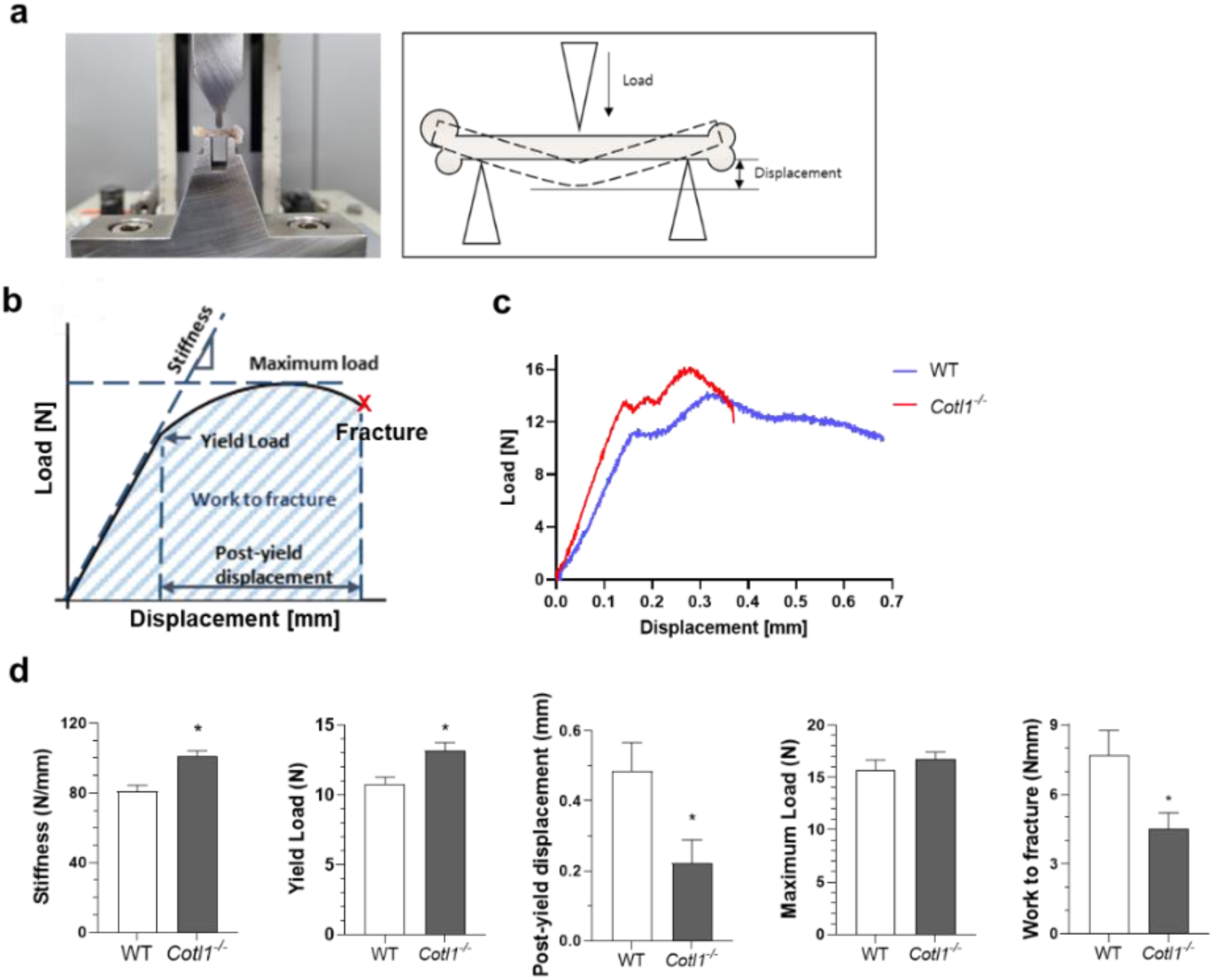
Three-point bending test in the femoral bone of wild-type (WT) and *Cotl1*^-/-^ mice. **a,** Three-point bending test device (left) and schematic view (right) of the experiment. **b,** Key outcome parameters of whole-bone mechanical properties. **c,** Load-displacement curves of WT and *Cotl1*^-/-^ mouse femurs under the three-point bending test. **d,** Quantification of the mechanical properties of WT and *Cotl1*^-/-^ mouse femurs, including stiffness (N/mm), yield load (N), post-yield displacement (mm), maximum load (N), and work to fracture (Nmm). \**p* < 0.05 vs. WT (Student’s *t*-test).

### Genetic association of SNPs in human *COTL1* gene with eBMD

To investigate whether *COTL1* gene is associated with the risk of bone-related traits, we conducted an association analysis of 6,047 SNPs in *COTL1* gene region for eBMD using the genotype and phenotype data (speed of sound measured by quantitative ultrasound) of 348,212 subjects from the UK Biobank^23^ (Figure S11). Association analysis was performed using PLINK software. Among the analyzed SNPs, rs2925049 showed the most significant association with eBMD (*p* = 2.84×10^−6^ and β = −0.0013), and its *p*-value was less than the Bonferroni-corrected threshold (*p*=8.26×10^−6^) (Figure S12). The regional plots of association analysis in the *COTL1* gene region revealed that several SNPs located near rs2925049 of *COTL1* showed a moderately significant association with eBMD, suggesting an important role of *COTL1* in bone density in humans.

## DISCUSSION

Osteopetrosis is a genetic disorder that causes the dysregulation of bone remodeling, resulting in increased BMD in the femur, pelvis, and skull^1^. Typical findings in patients with osteopetrosis include vulnerability to fractures, scoliosis, and sacroiliac joint pain^2^. In this study, we demonstrated that Cotl1 plays a fundamental role in osteoclastogenesis.

Firstly, a role of Cotl1 in platelet production in both zebrafish and mice (Figs. 1 and 2a). Platelets are closely associated with the regulation of bone remodeling^21^. Moreover, *in silico* pathway analysis of DEGs between the bone marrow of wild type and *Cotl1*^-/-^ mice suggested that Cotl1 may be involved in bone homeostasis (Fig 2b). Therefore, we focused on elucidating the involvement of Cotl1 in bone phenotypes. Primary-cultured mouse bone marrow-derived osteoclast lineage monocytes from *Cotl1*^-/-^ mice showed a decreased ability to differentiate into mature osteoclasts and impaired actin ring formation which is a critical process for osteoclast differentiation during bone remodeling (Fig 2). These findings suggest a fundamental role for Cotl1 in osteoclastogenesis. Structural and histological analyses of the femurs in *Cotl1*^-/-^ male and female mice revealed that loss of *Cotl1* results in sclerotic bone phenotypes (Figs. 3 and 4). In osteoporosis-induced female OVX mice, *Cotl1* knockout improves bone-related properties. Furthermore, mechanical assessment of *Cotl1*^-/-^ mouse femoral bone revealed a marble bone phenotype (the bones hardened and became denser), which was vulnerable to fractures (Fig. 5). Taken together, these results indicated that the loss of *Cotl1* results in osteopetrotic phenotypes through the inhibition of osteoclast differentiation.

HSC transplantation is the primary treatment for the most severe forms of osteopetrosis^1^. A previous study suggested a close correlation between bone metabolism and hematopoiesis^24^, and emerging evidence indicates that platelets are a unique blood element in the regulation of bone remodeling^25^. The main functions of platelets extend beyond hemostasis and wound healing; platelets further act as bone-healing blood factors and play an important role in repairing damaged bone structure^21^. Additionally, platelet size, volume, and BMD are positively correlated with bone-remodeling ability^25^. Remodeling of the bone marrow microenvironment and hematopoiesis are responses to the progression of immune thrombocytopenia, which is characterized by a reduction in platelet production^26^. Similarly, association studies have demonstrated a close relationship between platelet counts and bone mineralization, showing that high platelet counts are significantly correlated with osteopenia and osteoporosis^27^. Neonatal osteopetrosis presents with low platelet count associated with thrombocytopenia^28^. These studies suggest that platelets are among the major blood components associated with bone remodeling. Platelets can be detected by the expression of specific platelet membrane glycoproteins, such as CD41 and CD61, which are reliable molecular markers for the development of emerging HSCs^20^. Our study found that the predominant expression of *cotl1* in the caudal hematopoietic tissue of zebrafish and *cotl1* morphants was decreased expression of *c-myb*, a marker of HSCs, and CD41, a platelet receptor, suggesting that Cotl1 may be associated with bone remodeling.

Multipotent HSCs are responsible for platelet production. A previous study has shown that Cotl1 is a critical integrator of hemostasis and thrombus formation by modulating the biomechanical properties of platelets in mice^29^. The absence of *Cotl1* had no influence on platelet counts and function, but markedly affected platelet aggregate formation in 12- to 16-week old mice^29^. Similarly, in our study, no difference in platelet counts was found between both 18-week old WT and *Cotl1^-/-^* mice; however, a significant decrease in platelet production was observed in older mice aged 24-week old (Fig. 2a). Comparably, flow cytometry showed decreased expression of platelet surface markers (integrin α2b and β3) in the platelet-rich plasma of *Cotl1^-/-^* mice (Fig. 2a). Adding to the previous evidence that Cotl1 acts as a signal integrator for the regulation of thrombus formation in mouse platelets, our results suggest that Cotl1 contributes to platelet production in zebrafish and mice. Platelet function is involved in the maintenance of bone health (metabolism or hemostasis)^21^ and platelet activation, which is characterized by shape changes and enhanced osteoclastogenesis in bone marrow cells^30^. Multiple platelet formation and function processes are regulated by morphological changes that are mainly mediated by actin dynamics^31^. Additionally, actin dynamics play a critical role in the regulation of integrin signaling and platelet adhesion^31^. Integrin α_v_β_3_ is expressed in many types of cells, including osteoclasts, platelets, and megakaryocytes, and is a key factor for cell migration and invasion through the control of cytoskeletal organization, survival, and force generation^32^. Integrin α_v_β_3_ serves as a receptor for the extracellular matrix to mediate contact with the bone matrix, and the polarization of osteoclasts forms a unique actin structure such as a ruffled border and an actin ring to support the degradation of bone^33^. During osteoclast differentiation, integrin α_v_β_3_ production is increased, which binds to bone to support the resorption process^34^. A previous study demonstrated a strong connection between platelets and osteoclast β_3_ integrins in tumor-mediated bone destruction in osteoclast-defective *Src*-null mice^35^. Additionally, integrin β_3_-null mice showed dysfunctional aggregation of platelets and abnormal resorptive function of osteoclasts, resulting in a progressive osteosclerotic phenotype^36^. Our results showed that decreased levels of integrins, including integrin β_3_ (*Itgb3*) and integrin α_V_ (*Itgav*) in osteoclasts and integrin α2b (CD41) and integrin β3 (CD61) in platelets, which can promote the formation of dysfunctional osteoclasts and platelets, implying the osteopetrotic features in *Cotl1^-/-^* mice.

Osteopetrosis is mainly caused by malfunctioning osteoclast development^1^. The typical osteoclastogenesis pathway involves RANK signaling^5^ (Fig. 6). RANK, stimulated by RANKL binding, cooperates with integrin α_v_β_3_, stimulating the activation of c-Src and Pyk2 tyrosine kinases^37^. These events are associated with actin cytoskeleton-interacting proteins that regulate bone-resorbing osteoclast adhesion and migration^37^. The deletion of c-Src reduces osteoclastic resorptive function^38^, and mice genetically lacking *Pyk2* show impaired podosome organization and increased bone density^39^. Moreover, deficiency of actin cytoskeleton-interacting proteins, such as cofilin^15^, and gelsolin^16^ inhibits podosome belt assembly and formation, resulting in the disruption of osteoclast resorption. Our results showed that *Cotl1* absence resulted in abnormal actin ring formation, possibly through the downregulation of the integrins *Src*, *Ptk2b*, *Gsn*, and *Cfl1*. These results indicate that *Cotl1* contributes to actin ring formation in mature osteoclasts.

**Figure 6.**
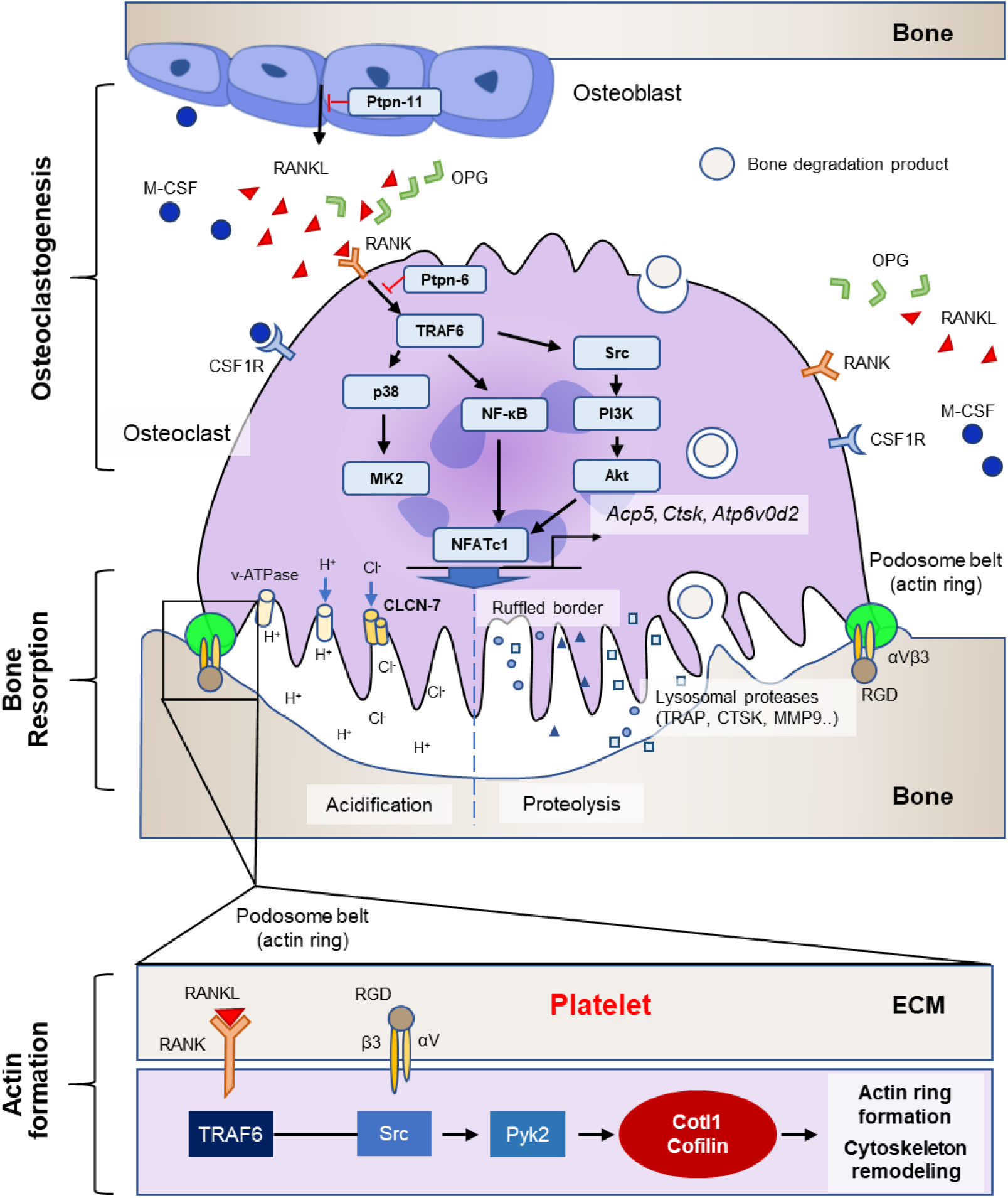
Schematic graphics of actin-binding protein-involved RANK signaling pathways for actin ring formation and osteoclastogenesis. RANK stimulated by RANKL binding cooperates with integrin αvβ3 to induce a canonical signaling pathway associated with actin cytoskeleton regulator proteins (Cotl1, Cofilin, etc.). Abbreviations: ECM, extracellular matrix; M-CSF, macrophage-colony stimulating factor; OPG, osteoprotegerin; RANK, receptor activator of NF-κB ligand; ANKL, RANK ligand; and RGD, Arginine-Glycine-Aspartic acid.

The osteoclastic podosome belt was positioned within the sealing zone and consisted of highly tyrosine-phosphorylated structures in osteoclasts^40^ (Fig. 6). Therefore, tyrosine phosphatases (PTPs), which regulate tyrosine phosphorylation, are primary regulators of osteoclast function and production^40^. The protein tyrosine phosphatases SHP-1 (encoded by *Ptpn-6*) and SHP-2 (encoded by *Ptpn-11*) are members of the Src homology 2 domain-containing cytoplasmic PTP family and are involved in osteoclast development^41^. SHP-2 is a well-known regulator of M-CSF- and RANK-evoked signaling, and *Shp2* mutant mice develop osteopetrosis with abnormal osteoclastic bone resorptive dynamics^42^, suggesting an important role of SHP-2 in bone homeostasis and osteoclastogenesis. However, SHP-1 negatively regulates osteoclast function and M-CSF signaling, which is a ligand of CSF1R^43^. We found that *Cotl1* loss reduced *Ptpn-11* expression without affecting *Ptpn-6* and *Csf1r* expression, and downregulated RANK signaling, suggesting that Cotl1 may be involved in the RANK-dependent pathway.

Upon binding of RANKL to RANK (encoded by *Tnfrsf11a*), the intracellular adapter TNF receptor-associated factor 6 (TRAF6) is recruited to activate signaling cascades, including NF-κB (NFKB1), MAP kinase-activated protein kinase 2 (MAPKAPK2), and AKT serine/threonine kinase 1 (AKT1), upregulating osteoclast-associated gene transcription via nuclear factor of activated T cells, cytoplasmic, calcineurin dependent 1 (NFATC1)^7, 44, 45^, the master transcription factor for osteoclast differentiation. NFATC1 can initiate the transcription of osteoclast-resorptive genes involved in acidification, such as ATPase H+ transporting V0 Subunit D2 (*Atpv0d2*) and chloride voltage-gated channel 7 (*Clcn7*), and proteolysis, such as TRAP (encoded by *Acp5*), cathepsin K (*Ctsk*), and matrix metalloproteinase-9 (*Mmp9*)^46, 47^ (Fig. 6). Our study revealed that *Cotl1* absence in osteoclasts promotes osteopetrosis by inhibiting platelet production, podosome formation, osteoclastogenesis, and osteoclast resorptive functions (acidification and proteolysis), which may be due to the downregulation of RANK cascades.

Bone remodeling is regulated by the balance between osteoblasts and osteoclasts, which works in concert with the secretion of several cytokines, including RANKL and OPG^5^. RANKL is an osteoclast differentiation factor that is generally secreted by activated T cells and osteoblasts and binds to RANK as an essential modulator of osteoclastogenesis and osteoclasts^6, 7^. OPG is an osteoclastogenesis inhibitory factor produced by bone marrow stromal cells and osteoblasts that prevents excessive bone resorption by acting as a decoy receptor for RANKL^48^. As variations in the plasma levels of RANKL and OPG disrupt bone homeostasis by altering bone mass, the ratio of OPG to RANKL has been used as a key indicator of bone mass^48^. In this study, *Cotl1*^-/-^ mice showed a significantly increased plasma OPG/RANK ratio, suggesting a predominance of high bone mass in *Cotl1*^-/-^ mice.

The main characteristic of *in vivo* models of osteopetrosis is an increase in bone density due to reduced osteoclastogenesis, which can lead to fractures and bone malformations^1, 3^. This study found a decreased number of TRAP-positive cells with indistinct resorptive pits and impaired trabecular parameters, such as BMD, BV/TV, Tb.Th, and Tb.Sp, in both *Cotl1*^-/-^ male and female mice. The OVX mouse model has been widely used in osteoporosis research because female hormones are some of the main factors that influence osteoclast and osteoblast differentiation^49^. Representative OVX phenotypes include marked bone loss due to increased bone resorption and decreased bone formation^49^. Our results showed that *Cotl1* inhibition inhibited osteoclastogenesis in the OVX-induced osteoporotic mouse model (Fig. 4). These data demonstrate that *Cotl1*^-/-^ mice display an osteopetrosis phenotype.

Thus far, there have been no reports on *COTL1* mutations in the Online Catalogue of Human Genes and Genetic Disorders (OMIM) database (https://www.omim.org) or the Human Gene Mutation Database (https://www.hgmd.cf.ac.uk). Meanwhile, a genome-wide association study (GWAS) of heel QUS in 426,824 white British subjects from the UK Biobank found 518 significant loci^50^. Among the 301 novel loci, the *COTL1* gene was also included. The intronic rs2925049 SNP of human *COTL1* gene was reported to be significantly associated with heel BMD (*p* = 5×10^-9^, β = 0.0108). We also performed an association analysis of 6,047 SNPs in *COTL1* gene region for eBMD in subjects from the UK Biobank^23^. Using subject filtration and phenotype QC, 348, 212 subjects were selected from the UK Biobank (Figure S11). Association results showed that the same rs2925049 SNP in *COTL1* gene was most significantly associated with eBMD (*p* = 2.84×10^−6^ and β = −0.0013), and several SNPs located near rs2925049 showed meaningful, significant association with eBMD (Figure S12). As the rs2925049 SNP is located in the intron region, this variation might be responsible for the difference in *COTL1* gene expression among the genotypes, suggesting that *COTL1* might play an important role in determining bone density.

## Limitations

This study has some limitations. First, a previous study reported that the absence of *Cotl1* had no influence on platelet counts in 12- to 16-week old mice^29^. Similarly, our hematological observation also showed no abnormalities in examined blood parameters in *Cotl1*^-/-^ mice at 7, 12, and 18-week of ages. However, a significant decrease in platelet production was found in *Cotl1*^-/-^ mice aged 24-week old (Fig. 2a; Figure S2c). It is difficult to explain the reason why the reduced platelet count is detected only in older *Cotl1*^-/-^ mice, despite occurrence of the augmented bone density in younger *Cotl1*^-/-^ mice (Figs. 3, 4). Second, although we found that an intronic rs2925049 SNP of human *COTL1* gene was significantly associated with eBMD in the UK Biobank subjects (Figures S12) and this SNP was reported to have a significant association with heel BMD in the same cohort^50^, there is a lack of replication studies in other ethnic cohorts.

## Conclusion and Perspectives

This is the first report on the role of *Cotl1* in osteoclastogenesis, demonstrating that its loss results in osteopetrotic phenotypes *in vitro* and *in vivo.* Taken together, our results indicate that the absence of *Cotl1* reduces platelet-mediated osteoclastogenesis and thereby causes osteopetrosis phenotypes in mice. *Cotl1* knockout mouse may be a useful animal model for osteopetrosis research. Thus, Cotl1 may be a promising target for novel therapeutic strategies for the treatment and/or prevention of impaired osteoclastogenesis-mediated bone diseases such as osteopetrosis and osteoporosis.

## Data Availability

All data associated with this study are available from the corresponding author on reasonable request.

## Author contributions

S.-Y.J. and E.P initiated the study. E.P., S.-H.Y., C.-G.L., S-H.L. and S-Y.C., performed experiments, H-S. J., H.G.W., J.E.L. and B.O., contributed to the bioinformatic data analysis. E.P. and H-S. J. wrote first draft. S.-Y.J. reviewed and edited.

## REFERENCES

1. Stark Z, Savarirayan R. Osteopetrosis. Orphanet J Rare Dis. 2009;4:5.

2. Gresky J, Sokiranski R, Witzmann F, Petiti E. The Oldest Case of Osteopetrosis in a Human Skeleton: Exploring the History of Rare Diseases. Lancet Diabetes Endocrinol. 2020;8:806–808.

3. Feng X, McDonald JM. Disorders of Bone Remodeling. Annu. Rev. Pathol. 2011;6:121–145.

4. Pereira M. et al. Common Signalling Pathways in Macrophage and Osteoclast Multinucleation. J. Cell Sci. 2018;131.

5. Roodman GD. Regulation of Osteoclast Differentiation. Ann. N. Y. Acad. Sci. 2006;1068:100–109.

6. Mun SH, Park PSU, Park-Min KH. The M-CSF Receptor in Osteoclasts and Beyond. Exp. Mol. Med. 2020;52:1239–1254.

7. Armstrong AP. et al. A RANK/TRAF6-Dependent Signal Transduction Pathway Is Essential for Osteoclast Cytoskeletal Organization and Resorptive Function. J. Biol. Chem. 2002;277:44347–44356.

8. Väänänen HK, Horton M. The Osteoclast Clear Zone Is a Specialized Cell-Extracellular Matrix Adhesion Structure. J. Cell. Sci. 1995;108:2729–2732.

9. Takito J, Inoue S, Nakamura M. The Sealing Zone in Osteoclasts: A Self-Organized Structure on the Bone. Int. J. Mol. Sci. 2018;19.

10. Baron R, Neff L, Louvard D, Courtoy PJ. Cell-Mediated Extracellular Acidification and Bone Resorption: Evidence for a Low pH in Resorbing Lacunae and Localization of a 100-kD Lysosomal Membrane Protein at the Osteoclast Ruffled Border. J. Cell Biol. 1985;101:2210–2222.

11. Everts V. et al. Degradation of Collagen in the Bone-Resorbing Compartment Underlying the Osteoclast Involves Both Cysteine-Proteinases and Matrix Metalloproteinases. J. Cell Physiol. 1992;150:221–231.

12. Schachtner H, Calaminus SD, Thomas SG, Machesky LM. Podosomes in Adhesion, Migration, Mechanosensing and Matrix Remodeling. Cytoskeleton (Hoboken). 2013;70:572–589.

13. Saltel F, Destaing O, Bard F, Eichert D, Jurdic P. Apatite-Mediated Actin Dynamics in Resorbing Osteoclasts. Mol. Biol. Cell. 2004;15:5231–5241.

14. Destaing O, Saltel F, Géminard JC, Jurdic P, Bard F. Podosomes Display Actin Turnover and Dynamic Self-Organization in Osteoclasts Expressing Actin-Green Fluorescent Protein. Mol. Biol. Cell. 2003;14:407–416.

15. Zalli D. et al. The Actin-Binding Protein Cofilin and Its Interaction With Cortactin Are Required for Podosome Patterning in Osteoclasts and Bone Resorption In Vivo and In Vitro. J. Bone. Miner. Res. 2016;31:1701–1712.

16. Chellaiah M. et al. Gelsolin Deficiency Blocks Podosome Assembly and Produces Increased Bone Mass and Strength. J. Cell Biol. 2000;148:665–678.

17. Poukkula M, Kremneva E, Serlachius M, Lappalainen P. Actin-Depolymerizing Factor Homology Domain: a Conserved Fold Performing Diverse Roles in Cytoskeletal Dynamics. Cytoskeleton (Hoboken). 2011;68:471–490.

18. Hou X, Nozumi M, Nakamura H, Igarashi M, Sugiyama S. Coactosin Promotes F-Actin Protrusion in Growth Cones Under Cofilin-Related Signaling Pathway. Front. Cell Dev. Biol. 2021;9:660349.

19. Bolger-Munro M. et al. The Wdr1-LIMK-Cofilin Axis Controls B Cell Antigen Receptor-Induced Actin Remodeling and Signaling at the Immune Synapse. Front. Cell Dev. Biol. 2021;9:649433.

20. Mateo A, Perez de la Lastra JM, Garrido JJ, Llanes D. Platelet Activation Studies with Anti-CD41/61 Monoclonal Antibodies. Vet Immunol Immunopathol. 1996;52:357–362.

21. Sharif PS, Abdollahi M. The Role of Platelets in Bone Remodeling. Inflamm. Allergy Drug. Targets. 2010;9:393–399.

22. Jepsen KJ, Silva MJ, Vashishth D, Guo XE, van der Meulen MC. Establishing Biomechanical Mechanisms in Mouse Models: Practical Guidelines for Systematically Evaluating Phenotypic Changes in the Diaphyses of Long Bones. J. Bone Miner. Res. 2015;30:951–966.

23. Sudlow C., et al. UK Biobank: an Open Access Resource for Identifying the Causes of a Wide Range of Complex Diseases of Middle and Old Age. PLoS Med. 2015;12:e1001779.

24. Suda T. Hematopoiesis and Bone Remodeling. Blood. 2011;117:5556–5557.

25. Salamanna F, Maglio M, Sartori M, Tschon M, Fini M. Platelet Features and Derivatives in Osteoporosis: A Rational and Systematic Review on the Best Evidence. Int. J. Mol. Sci. 2020;21.

26. Herd OJ. et al. Bone Marrow Remodeling Supports Hematopoiesis in Response to Immune Thrombocytopenia Progression in Mice. Blood Adv. 2021;5:4877–4889.

27. Kim J, Kim HS, Lee HS, Kwon YJ. The Relationship between Platelet Count and Bone Mineral Density: Results from Two Independent Population-Based Studies. Arch. Osteoporos. 2020;15:43.

28. Kazemian M, Fallahi M, Fakhraee SH, Alavi S. Osteopetrosis Presenting with Neonatal Thrombocytopenia: A Case Report. Iran. J. Neonatol. 2018;9.

29. Scheller I. et al. Coactosin-Like 1 Integrates Signaling Critical for Shear-Dependent Thrombus Formation in Mouse Platelets. Haematologica. 2020;105:1667–1676.

30. Weicht B. et al. Activated Platelets Positively Regulate RANKL-Mediated Osteoclast Differentiation. J Cell Biochem. 2007;102:1300–1307.

31. Bearer EL, Prakash JM, Li Z. Actin Dynamics in Platelets. Int. Rev. Cytol. 2002;217:137–182.

32. Hood JD, Cheresh DA. Role of Integrins in Cell Invasion and Migration. Nat. Rev. Cancer. 2002;2:91–100.

33. Nakamura I. et al. Role of alpha(v)Beta(3) Integrin in Osteoclast Migration and Formation of the Sealing Zone. J. Cell Sci. 1999;112:3985–3993.

34. Engleman VW. et al. A Peptidomimetic Antagonist of the alpha(v)beta3 Integrin Inhibits Bone Resorption In Vitro and Prevents Osteoporosis In Vivo. J. Clin. Invest. 1997;99:2284–2292.

35. Bakewell SJ. et al. Platelet and Osteoclast beta3 Integrins Are Critical for Bone Metastasis. Proc. Natl. Acad. Sci. U. S. A. 2003;100:14205–14210.

36. McHugh KP. et al. Mice Lacking beta3 Integrins Are Osteosclerotic Because Of Dysfunctional Osteoclasts. J. Clin. Invest. 2000;105:433–440.

37. Sanjay A. et al. Cbl Associates with Pyk2 and Src to Regulate Src Kinase Activity, alpha(v)Beta(3) Integrin-Mediated Signaling, Cell Adhesion, and Osteoclast Motility. J. Cell Biol. 2001;152:181–195.

38. Miyazaki T, Tanaka S, Sanjay A, Baron R. The Role of c-Src Kinase in the Regulation of Osteoclast Function. Mod. Rheumatol. 2006;16:68–74.

39. Gil-Henn H. et al. Defective Microtubule-Dependent Podosome Organization in Osteoclasts Leads to Increased Bone Density in Pyk2(-/-) Mice. Journal of Cell Biol. 2007;178:1053–1064.

40. Shalev M, Elson A. The Roles of Protein Tyrosine Phosphatases in Bone-Resorbing Osteoclasts. Biochim. Biophys. Acta. Mol. Cell Res. 2019;1866:114–123.

41. Yang H, Wang L, Shigley C, Yang W. Protein Tyrosine Phosphatases in Skeletal Development and Diseases. Bone Res. 2022;10:10.

42. Zhou Y. et al. SHP2 Regulates Osteoclastogenesis by Promoting Preosteoclast Fusion. FASEB J. 2015;29:1635–1645.

43. Umeda S. et al. Deficiency of SHP-1 Protein-Tyrosine Phosphatase Activity Results in Heightened Osteoclast Function and Decreased Bone Density. Am. J. Pathol. 1999;155:223–233.

44. Abu-Amer Y. NF-kappaB Signaling and Bone Resorption. Osteoporos. Int. 2013;24:2377–2386.

45. Wong BR. et al. TRANCE, a TNF Family Member, Activates Akt/PKB through a Signaling Complex Involving TRAF6 and c-Src. Mol. Cell. 1999;4:1041–1049.

46. Kim K, Lee SH, Ha Kim J, Choi Y, Kim N. NFATc1 Induces Osteoclast Fusion via Up-Regulation of Atp6v0d2 and the Dendritic Cell-Specific Transmembrane Protein (DC-STAMP). Molecular Endocrinology. 2008;22:176–185.

47. Takayanagi H. et al. Induction and Activation of the Transcription Factor NFATc1 (NFAT2) Integrate RANKL Signaling in Terminal Differentiation of Osteoclasts. Dev. Cell. 2002;3:889–901.

48. Tanaka H, Mine T, Ogasa H, Taguchi T, Liang CT. Expression of RANKL/OPG during Bone Remodeling In Vivo. Biochem. Biophys. Res. Commun. 2011;411:690–694.

49. Khosla S, Oursler MJ, Monroe DG. Estrogen and the Skeleton. Trends Endocrinol. Metab. 2012;23:576–581.

50. Morris JA. et al. An Atlas of Genetic Influences on Osteoporosis in Humans and Mice. Nat. Genet. 2019;51:258–266.

